# Exome Sequencing of 963 Chinese Families Identifies Novel Epilepsy Candidate Genes

**DOI:** 10.1101/2024.06.11.24308522

**Authors:** Yu Ma, Ke Su, Mingshan Zhou, Yihan Liu, Guangqin Lu, Jie Wang, Chengjie Li, Tianqi Wang, Yingfeng Li, Qi Zhang, Xia Guan, Xiong Han, Wenling Li, Rongna Ren, Min Zhong, Ding Ding, Yonghui Jiang, Gang Peng, Yi Wang, Qihui Wu, Shaohua Fan

**Affiliations:** Department of Neurology, Children’s Hospital of Fudan University, State Key Laboratory of Genetic Engineering, Human Phenome Institute and School of Life Sciences, Lab for Evolutionary Synthesis, Institute of Brain Science, State Key Laboratory of Medical Neurobiology, MOE Frontiers Center for Brain Science, Institutes of Brain Science, Department of Neurology, Huashan Hospital of Fudan University, Fudan University, Shanghai 200433, China; Shanghai Key Laboratory of Anesthesiology and Brain Functional Modulation, Clinical Research Center for Anesthesiology and Perioperative Medicine, Translational Research Institute of Brain and Brain-Like Intelligence, Shanghai Fourth People’s Hospital Affiliated to Tongji University School of Medicine, State Key Laboratory of Cardiology and Medical Innovation Center, Shanghai East Hospital, School of Medicine, Tongji University, Shanghai 200434, China; Department of Neurology, Henan Provincial People’s Hospital, Henan Province, Zhengzhou 450003, China; Neurosurgery Department of Epilepsy, the Second Hospital of Hebei Medical University, Shijiazhuang 050000, China; Department of Pediatrics, General Hospital of Fuzhou, Military Command, Fujian Medical University, Fuzhou 350025, China; Department of Neurology, Children’s Hospital of Chongqing Medical University, Chongqing 400014, China; Department of Genetics, Neuroscience, and Pediatrics, Yale University School of Medicine, New Haven CT 06510, USA

## Abstract

Epilepsy, a prevalent neurodevelopmental disorder in children, is often accompanied by detrimental psychological consequences and other comorbidities. We performed exome sequencing on 963 patient-parent trios, revealing differences in genetic epidemiology between Chinese and European epilepsy cohorts. The diagnostic yield for known epilepsy genes was 40%. Pathogenic variants were most commonly found in SCN1A, KCNQ2, and DEPDC5. Additionally, we identified 15 novel monogenic epilepsy candidates in at least two patients diagnosed with developmental and epileptic encephalopathy, non-acquired focal epilepsy, or genetic generalized epilepsy, including *ADCY2, BCAR3, CDC45, CHRNG, CRTC2, CSMD1, CSMD2, KDM6B, KIF1B, PLEKHM3, PPP4R1, RASGRP2, SGSM2, SYNE1*, and *ZFHX3*. Aside from *ADCY2*, which was implicated in the GABAergic synapse pathway based on KEGG analysis, these candidates do not belong to known epilepsy pathways. Local field potential recordings in zebrafish and calcium imaging experiments validated associations for 11 of these genes, excluding those unsuitable for functional analyses. Furthermore, we found that *CRTC2* overexpression leads to hippocampal neuronal hyperactivity using multi-electrode arrays and electrophysiology. We have documented the first-line medications prescribed for patients harboring variants in the novel candidate genes. This study expands our understanding of the genetic underpinnings of epilepsy and provides opportunities for early diagnosis and personalized medicine approaches.

## Introduction

Children with epilepsy diagnosed before the age of three years are at increased risk for cognitive and behavioral difficulties^1–3^. This risk is compounded by drug-resistant seizures and a high frequency of seizures^1–3^. Therefore, the concept of developmental and epileptic encephalopathy (DEE) was introduced to describe children with early-onset drug-resistant epilepsy and encephalopathy, which can cause developmental delays or regression^4,5^. The other two major types of epilepsy include genetic generalized epilepsy (GGE) and non-acquired focal epilepsy (NAFE), which is characterized by seizures involving both hemispheres of the brain and localized cortical region^6^, respectively.

The development of epilepsy is complex, with genetic factors playing a substantial role. Early twin and family-based studies indicate genetics underlie approximately 70 to 80% of epilepsy cases, particularly those presenting in neonates and children^7–9^. Recent advancements in next-generation sequencing (NGS) technologies, notably the advent of exome sequencing (ES), have revolutionized the etiological diagnosis of epilepsy^6,10–15^. Until 2023, more than 900 monogenic “epilepsy genes” had been identified and ∼90% of them were associated with DEE^16^. In contrast, around 5% of all monogenic epilepsy genes were associated with a GGE and/or NAFE phenotype^16^. Previous genome-wide association studies (GWAS) also provide evidence for oligogenic or polygenic causes of GGE and NAFE^17–21^.

Despite a large number of genes or loci being identified in previous studies, our understanding of the genetic underpinnings of epilepsy remains exclusive. For example, diagnostic rates in sequencing-based studies have ranged from 18% to 48% due to case selection variances, sequencing methodologies (such as capture-based sequencing, whole genome sequencing, or exome sequencing), the number of genes examined, and cohort sizes^6,10–14^. On the other hand, the cumulative effects of all associated loci identified by epilepsy GWAS can explain only a fraction of the estimated heritability from family studies^17–21^. Furthermore, most large-scale epilepsy genomic investigations to date, including seminal efforts by the Epi4K and Epi25K consortia, have concentrated predominantly on individuals of European ancestry^6,10,22–27^ although some studies have studied samples of other ancestral backgrounds^9,14,28,29^. Therefore, our understanding of the genetic basis of epilepsy is still exclusive.

In this study, we leveraged ES of 963 Chinese proband-parent trios, combined with granular clinical phenotyping, to investigate the genetic architecture of epilepsy. With a cohort size almost two times larger than previous efforts in this population^30^, our approach enabled accurate delineation of the prevalence and mutational spectra of established epilepsy genes while uncovering novel monogenic candidate genes observed recurrently (in at least two patients). We conducted extensive functional validations in zebrafish and cellular models for the candidate genes. By reviewing patient medication history and electronic health records, we also identified the first-line anti-seizure medications for patients harboring variants in either known or novel epilepsy genes. Our integrated strategy promises improved genomic diagnosis rates, facilitating early detection and personalized therapeutic decisions.

## Results

### Demographics and clinical data

The present study recruited a total of 963 children with epilepsy. Consistent with previous studies^31–33^, we observed a higher incidence of epilepsy in males (573, 59.5%) compared to females (390, 40.5%), with a male-to-female ratio of 1.47 to 1. The mean reported age of seizure onset was 40.5 months (95% confidence interval 38.0-43.0, with the highest incidence (306, 31.8%) occurring in children in their infancy stage (between one month to one year) compared to other stages. Focal onset (654, 67.9%) was the most common onset type of seizure, including focal to bilateral type (274/654, 41.9%), followed by generalized onset (291, 30.2%), and unknown onset type (18, 1.9%) (**Fig. 1A**). Furthermore, we observed that 87.9% of generalized seizure onset cases were motor seizures, with the tonic-clonic subtype (36.1%) being the most frequent, followed by spasm (14.1%) and tonic seizures (11.3%). For patients with no obvious seizure phenotype (non-motor seizures), 9.6% and 2.4% of patients exhibited typical absence seizures and atypical absence seizures, respectively. For patients with an unknown seizure onset, motor seizure accounted for 27.8% and the remaining 72.2% were of unknown type (**Fig. 1A**). Finally, consistent with previous studies^29,34^, 175 (18.2%) of patients exhibited more than two types of seizures (**Fig. 1A**).

**Fig. 1:**
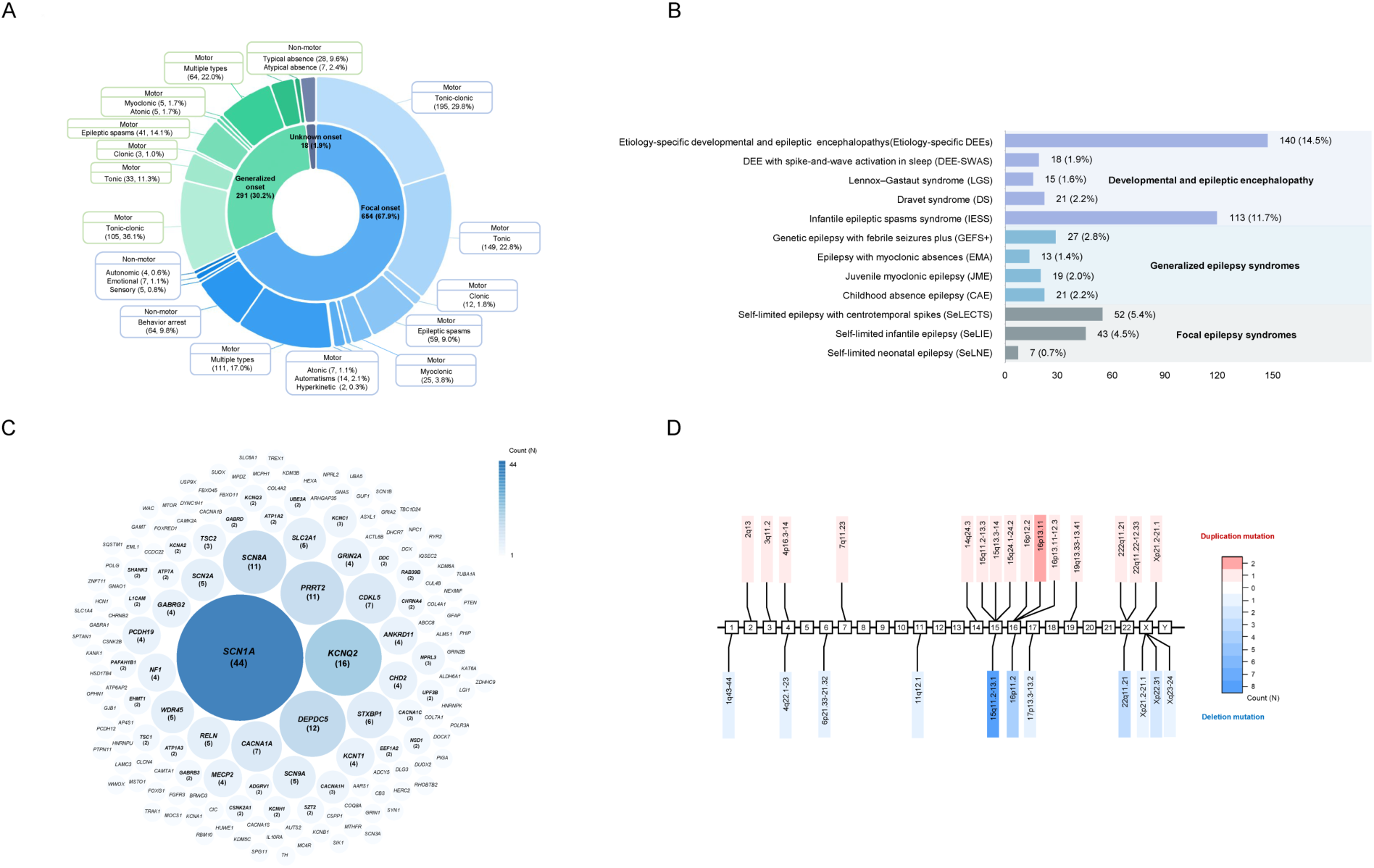
Overview of seizure and syndrome classification of 963 Patients with epilepsy in the present study. A: Summary of the seizure types of 963 patients in the present study. We classified the seizure types based on the International League Against Epilepsy Criteria (2017). B: The classifications of epilepsy syndromes of 489 patients, the rest 474 (49.2%, 474 out of 963) of the patients exhibit no explicit epileptic syndrome based on the International League Against Epilepsy Criteria classification. C: The numbers of patients with pathogenic/likely pathogenic mutations in known epilepsy genes. The size of the circle and the numbers in the bracket represent the number of mutations per gene. The circles without numbers indicate that only one mutation was identified in the gene. D: Genomic locations of 26 pathogenic/likely pathogenic CNVs that were detected in 40 patients. The numbers in the box indicate the chromosome names. The color bar shows the number of copy number variations, blue: deletion, red: duplication.

Of the 963 patients studied, 422 (43.8%) were diagnosed with DEE, 140 (14.5%) were diagnosed with genetic generalized epilepsy (GGE), 389 (40.4%) were diagnosed with non-acquired focal epilepsy (NAFE), and 12 (1.3%) were unclassified. Common epileptic syndromes were observed in 50.8% of the patients. Patients with focal epilepsy syndromes were diagnosed as self-limited neonatal epilepsy (SeLNE) in 7 cases (0.7%), self-limited infantile epilepsy (SeLIE) in 43 cases (4.5%), and self-limited epilepsy with centrotemporal spikes (SeLECTS) in 52 cases (5.4%) **(Fig. 1B)**. In addition, we observed that the patients with generalized epilepsy syndromes were childhood absence epilepsy (CAE) (21, 2.2%), juvenile myoclonic epilepsy (JME) (19, 2.0%), epilepsy with myoclonic absences (EMA) (13, 1.4%) and genetic epilepsy with febrile seizures plus (GEFS+) (27, 2.8%) **(Fig. 1B)**. Furthermore, patients with DEE were diagnosed with infantile epileptic spasms syndrome (IESS) (113, 11.7%), Dravet syndrome (DS) (21, 2.2%), Lennox– Gastaut syndrome (LGS) (15, 1.6%), DEE with spike-and-wave activation in sleep (DEE-SWAS) (18, 1.9%), and Etiology-specific DEEs (140, 14.5%) **(Fig. 1B)**. The remaining ∼49.2% (474/963) of the patients had no explicit epileptic syndrome classification.

We also observed that a significant proportion of patients demonstrated neuropsychiatric comorbidities. Specifically, 43.8% of patients had intellectual disabilities, 13.0% had attention deficit hyperactivity disorder (ADHD), and 4.6% had autism spectrum disorder (ASD). Additionally, 37.7% of the patients are intractable. Based on brain MRI, we discovered epileptogenic abnormalities, including brain malformations, dysplasia, and atrophy, in 525 (54.5%) of the patients. **Supplementary Table 1** contains a detailed overview of the demographic and clinical characteristics of the patients.

### Genetic diagnoses based on clinical testing

In the present study, we first examined 963 patients’ clinical testing results based on exome sequencing (ES). Based on the ES results, 39.6% (381/963) of patients received positive clinical diagnoses. We identified 329 pathogenic or likely pathogenic single nucleotide variants (SNVs) in 159 known epilepsy genes across 341 patients (**Fig. 1C**), following the American College of Medical Genetics (ACMG) guidelines. Missense variants were the most frequent variant type (210, 55.1%), followed by insertions and deletions (indels) (75, 19.7%), nonsense (36, 9.5%), and splicing variants (20, 5.2%). Among the known epilepsy genes, *SCN1A* harbored the greatest number of pathogenic/likely pathogenic variants (44, 12.9%, of which 20 were novel), followed by *KCNQ2* (16, 4.7%, of which 5 were novel) and *DEPDC5* (12, 3.5%, of which 5 were novel). *SCN1A, KCNQ2*, and *DEPDC5* are associated with Dravet syndrome or generalized epilepsy with febrile seizures plus 2 (GEFS+2), DEE7 or benign familial neonatal seizures (BFNS1), and familial focal epilepsy with variable foci-1 (FFEVF1), respectively. Additionally, 26 CNVs, ranging in size from 607 Kb to 7.66 Mb, were detected in 40 patients. 15q11.2-13.1 deletions (8, 20%) were the most common, followed by 16p deletions (4, 10%) and 16p duplications (4, 10%) (**Fig. 1D**).

Of the 381 patients with pathogenic or likely pathogenic variants, the median age of seizure onset was 11 months, with the highest incidence (n = 179, 47.0%) occurring during infancy. By epilepsy type, 236 (61.9%) patients were diagnosed with DEE, 45 (11.8%) with GGE, 95 (24.9%) with NAFE, and 5 (1.4%) were unclassified.

We noted that earlier seizure onset age correlated with higher rates of genetic diagnosis. The rate of genetic diagnosis was highest during the neonatal period at 75.0%, followed by infancy at 58.5%, and toddlerhood at 38.7%. Furthermore, the rate of genetic diagnosis also varied by epilepsy type. DEE had the highest rate at 55.9% (236/422), followed by GGE at 32.1% (45/140), and NAFE at 24.4% (95/389).

### A genetic epidemiology difference between Chinese and European epilepsy cohorts

When comparing the top 15 genes harboring the highest burden of pathogenic or likely pathogenic variants between our cohort and two other European cohorts^24,35^, some genes consistently emerge, including *SCN1A* and *STXBP1*, which consistently rank among the top 15 genes across various studies. However, discrepancies exist between samples of Chinese and European ancestries (**Figs. 2A-C**). For example, although *PRRT2* ranks fourth in our cohort in terms of the highest-burden of pathogenic or likely pathogenic mutations, it does not feature among the top 15 genes in the other two European cohorts (**Figs. 2A-C**). Some genes, such as *GABRB3* and *DNM1*, are among the top 15 genes with the highest burden of variants in European cohorts but were not ranked as the top 15 genes in our cohort **(Figs. 2A-C)**.

**Fig. 2:**
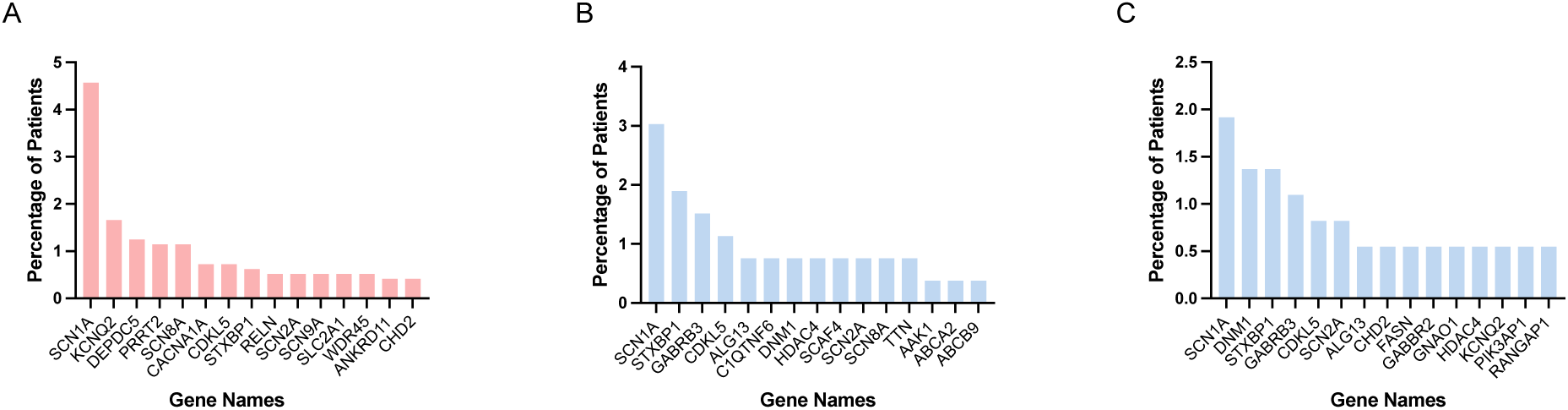
A genetic epidemiology difference between Chinese and European epilepsy cohorts. A-C: The genetic epidemiology difference between Chinese and European epilepsy cohorts. We compared the top 15 genes harboring the highest burden of pathogenic or likely pathogenic variants between our cohort (A) and two other European cohorts, EPGP (B) and EuroEPINOMICS and Epi4K/EPGP (C) ^24,35^. The X- and Y-axis indicates gene names and the percentage of patients in the cohort, respectively.

### Novel candidate genes for epilepsy

Aiming for the identification of novel candidate genes for epilepsy, we analyzed the ES data from 577 patients with previously undiagnosed epilepsy and their parents (the core family cohort), as well as variant calling results from 5 additional patients whose raw sequencing data were unavailable. Of the 582 patients, 186 (32.0%) were classified as having DEE, 95 (16.3%) with GGE, 294 (50.5%) with NAFE, and seven patients were unclassifiable (**Supplementary Table 1**).

We performed joint variant calling using the GATK toolkit following the germline variant calling best practices and used all the variants with the “PASS” flag in the downstream analysis. The functional impact of the variants was predicted using ANNOVAR based on nine in silico pathogenicity prediction programs. A variant is defined as “pathogenic” if it causes gene disruption (e.g., mutations at splicing sites, pre-stop codon gain and stop codon loss, start codon loss, open reading frame shift) or damaging missense effect (predicted to have deleterious effects by at least 3 in silico programs or with CADD score greater than or equal to 20 or DANN score greater than or equal to 0.93) and with frequency ≤ 0.001 in the EAS population of gnomAD, 1000 Genomes Project, and ExAc databases. We retained *de novo* “pathogenic” variants and inherited “pathogenic” variants and assessed whether these variants were not located in the known epilepsy genes in the Online Mendelian Inheritance in Man (OMIM, https://www.omim.org/), HGMD^36^, Clinvar^37^, and PubMed databases. We defined the genes that contained variants passing our filtering and interpretation procedures as “novel candidate genes of epilepsy”. It is worth noting that we prioritized candidate genes that were identified in at least two unrelated probands.

Our analysis identified 2,343,856 total variants in the patients, including 2,869 de novo variants (1,757 SNVs and 1,112 indels) observed in 555 patients, and 2,340,987 inherited variants (comprising 2,190,580 SNVs and 150,407 indels) detected in 577 patients. On average, there were around 5 *de novo* variants per patient (range 1-29) and ∼4,050 inherited variants per patient (range 3,641-4,889). We inferred that 17.29% (496 out of 2,869) *de novo* variants and 4.48% (104,877 out of 2,340,987) inherited variants were likely pathogenic, defined as variants causing gene disruption (such as splicing site, stop gain/loss, start codon loss, frameshift) or damaging missense variants (predicted deleterious by ≥ 3 *in silico* pathogenicity prediction programs and allele frequency ≤ 0.001 in East Asian populations of gnomAD, 1000 Genomes, and ExAC databases). On average, we identified around 0.86 likely pathogenic *de novo* and 181 inherited variants per patient.

Our analysis indicates that 53 pathogenic variants in 17 genes (*ABCA2*, *ADCY2*, *BCAR3*, *CDC45*, *CHRNG*, *CRTC2*, *CSMD1*, *CSMD2*, *KDM6B*, *KIF1B*, *LAMA5*, *PLEKHM3*, *PPP4R1*, *RASGRP2*, *SGSM2, SYNE1,* and *ZFHX3*) were identified in at least two patients but not in their healthy relatives. This provides a potential genetic cause for 78 patients (including 41 probands and 37 relatives) of 48 families (including one proband deep-exome-sequencing), which accounts for 7.04% (41/582) of the patients with negative clinical genetic testing. Among these 17 genes, *ABCA2*^38^ and *LAMA5*^39^ are known monogenic epilepsy genes and the rest 15 genes have not been reported to play a role in epilepsy based on the searches of the Online Mendelian Inheritance in Man (OMIM) as of Dec 12 2023 and Genes4Epilepsy^16^ databases (EpilepsyGenes_v2023-09.tsv). Besides, we noted that none of the 15 candidates is significant in studies between 9,170 epilepsy-affected individuals and 8,436 controls^6^ and 20,979 epilepsy cases and 33,444 controls^15^ based on burden test (**Supplementary Table 2**). In addition, no damaging missense variants present in the 20,979 cases with epilepsy analyzed by the Epi25 Collaborative (**Supplementary Table 2**). Therefore, we defined these 15 genes as novel monogenic epilepsy candidates.

We observed that *BCAR3* and *CSMD2* harbor missense and frameshift variants, *CRTC2* harbors missense and stop-gain variants, and the rest 12 genes carry only missense variants. According to the Simons Foundation Autism Research Initiative gene scoring scheme^40^, ten (*BCAR3, CDC45*, *CRTC2*, *CSMD1*, *CSMD2, PPP4R1, RASGRP2, SGSM2*, *SYNE1*, and *ZFHX3*) and five genes (*ADCY2*, *CHRNG*, *KDM6B*, *KIF1B*, and *PLEKHM3*) can be classified as “high confidence” and “strong candidates,” respectively. This is based on the presence of pathogenic variants in at least three unrelated patients for the former and at least two unrelated patients for the latter.

Based on the gene annotations in the OMIM database, the 15 novel candidates are involved in diverse functional domains, including synaptic function (*CHRNG*), intracellular cytoskeleton (*KIF1B* and *SYNE1*), extracellular fibronectin (*CSMD1* and *CSMD2*), intracellular signaling transduction (*ADCY2, BCAR3, PLEKHM3, PPP4R1, RASGRP2* and *SGSM2*), and DNA duplication (*CDC45*) as well as transcription regulation or epigenetic modification (*CRTC2, KDM6B*, and *ZFHX3*) in cellular nucleus **(Fig. 3A)**.

**Fig. 3:**
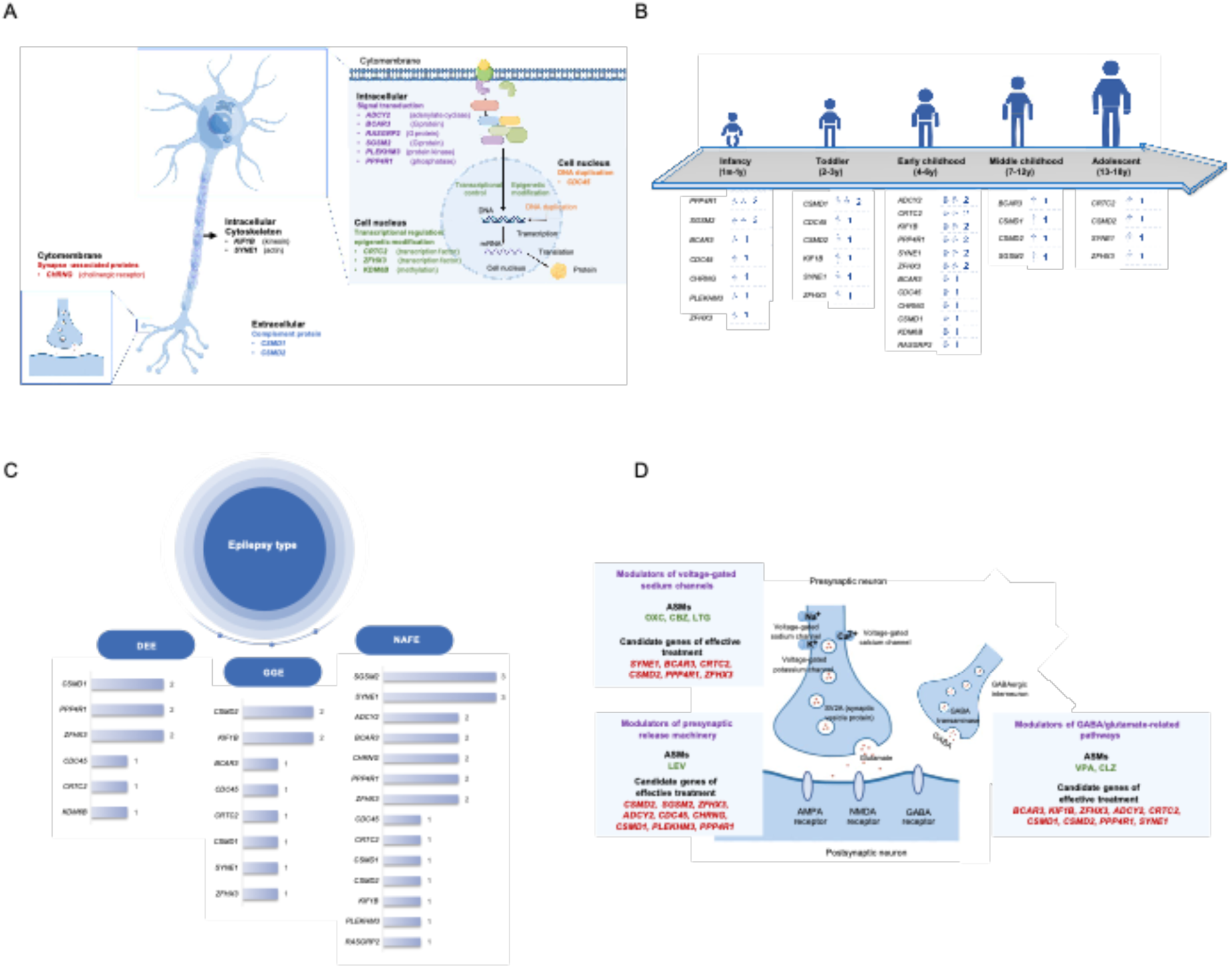
Functional domains of 15 novel epilepsy candidate genes and associated Clinical Features. A: The prospective functional domains of 15 novel epilepsy candidates. We assigned the functional domains of the candidates based on the OMIM database (https://omim.org/). B: Age of seizure onset of probands with the variants in the 15 novel epilepsy candidate genes. The numbers following the gene names are the number of patients. m: month, y: year C: Types of epilepsy diagnosed in the patients carrying the variants in the novel epilepsy candidate genes. The number on the bar indicates the number of patients per gene. DEE: developmental and epileptic encephalopathy, GGE: genetic generalized epilepsy, NAFE: non-acquired focal epilepsy. D: Potential effective anti-seizure medications (ASMs) for the novel epilepsy candidate genes. We observed that 76.5% (26 of 34) of the patients’ seizures were controlled using ASMs that targeted a wide range of pathways, including the modulators of presynaptic release machinery (e.g., LEV), modulators of voltage-gated sodium channels (e.g., OXC, CBZ, and LTG), and modulators of GABA/glutamate-related pathways (e.g., VPA and CLZ).

To further understand the functional pathways of the 15 candidate genes, we first merged them with the 978 previously reported epilepsy genes in the Genes4Epilepsy databases. As 15 genes would be insufficient for statistically significant pathway enrichment analysis, expanding the set provided more power to detect enriched pathways. We then performed a KEGG enrichment test using DAVID^41^. Our findings revealed that the majority of the identified genes were not associated with canonical epilepsy pathways, including mTOR, GABAergic synapse, and potassium/calcium channel pathways. Notably, *ADCY2* emerged as an exception, being implicated in the GABAergic synapse pathway **(Supplementary Table 3)**.

Based on their electronic health records, 34 probands harboring pathogenic variants in the novel epilepsy candidate genes displayed diverse ages of seizure onset, ranging from 3 months to 15 years **(Fig. 3B),** and types of epilepsy, including DEE, GGE, and NAFE **(Fig. 3C)**. Regarding epilepsy prognosis and drug prescription, 26 of 34 (76.5%) probands achieved seizure control using anti-seizure medications (ASMs) targeting diverse pathways. For instance, we observed that modulators of presynaptic release machinery (e.g. levetiracetam) effectively treated patients with variants in *CSMD2*, *SGSM2*, *ZFHX3*, *ADCY2*, *CDC45*, *CHRNG*, *CSMD1*, *PLEKHM3*, and *PPP4R1*. Voltage-gated sodium channel modulators (e.g. oxcarbazepine, carbamazepine, lamotrigine) benefited patients with variants in *SYNE1*, *BCAR3*, *CRTC2*, *CSMD2*, *PPP4R1*, and *ZFHX3*. Patients with variants in *BCAR3*, *KIF1B*, *ZFHX3*, *ADCY2*, *CRTC2*, *CSMD1*, *CSMD2*, *PPP4R1*, and *SYNE1* responded to modulators of GABA/glutamate pathways (e.g. valproic acid, clonazepam) (**Fig. 3D**).

We also investigated the potential functional impacts of the *de novo* pathogenic variants that were identified in one patient. When focusing on 502 *de novo* variants in 382 autosomal genes from 105 patients with DEE, 261 *de novo* variants in 198 genes in 55 patients with GGE, and 809 *de novo* variants in 643 genes in 147 patients with NAFE, we observed these genes were significantly enriched (adjusted *P*-value <0.01) in pathways involving in cell communication, response to stimulus, signal transduction, and neuron development based on a gene-based GO enrichment test using the online version of g:Profiler^42^ (**Supplementary Table 4**).

### Examine the prospective functional impacts of the candidates based on Cas9- mediated gene knockout in F0 zebrafish

We generated biallelic knockouts in F0 zebrafish using the CRISPR-Cas9 method^43^ to explore the functional impacts of the novel candidates. The impacts of knockouts on brain activities were monitored by local field potential (LFP) recordings from the forebrain regions in larval zebrafish^44^ (**Fig. 4A**). To validate the system, we knocked out *BSCL2*^45,46^, which is a known epilepsy gene, in F0 zebrafish and observed a significant increase in synchronized epileptiform discharge compared with the controls.

**Fig. 4:**
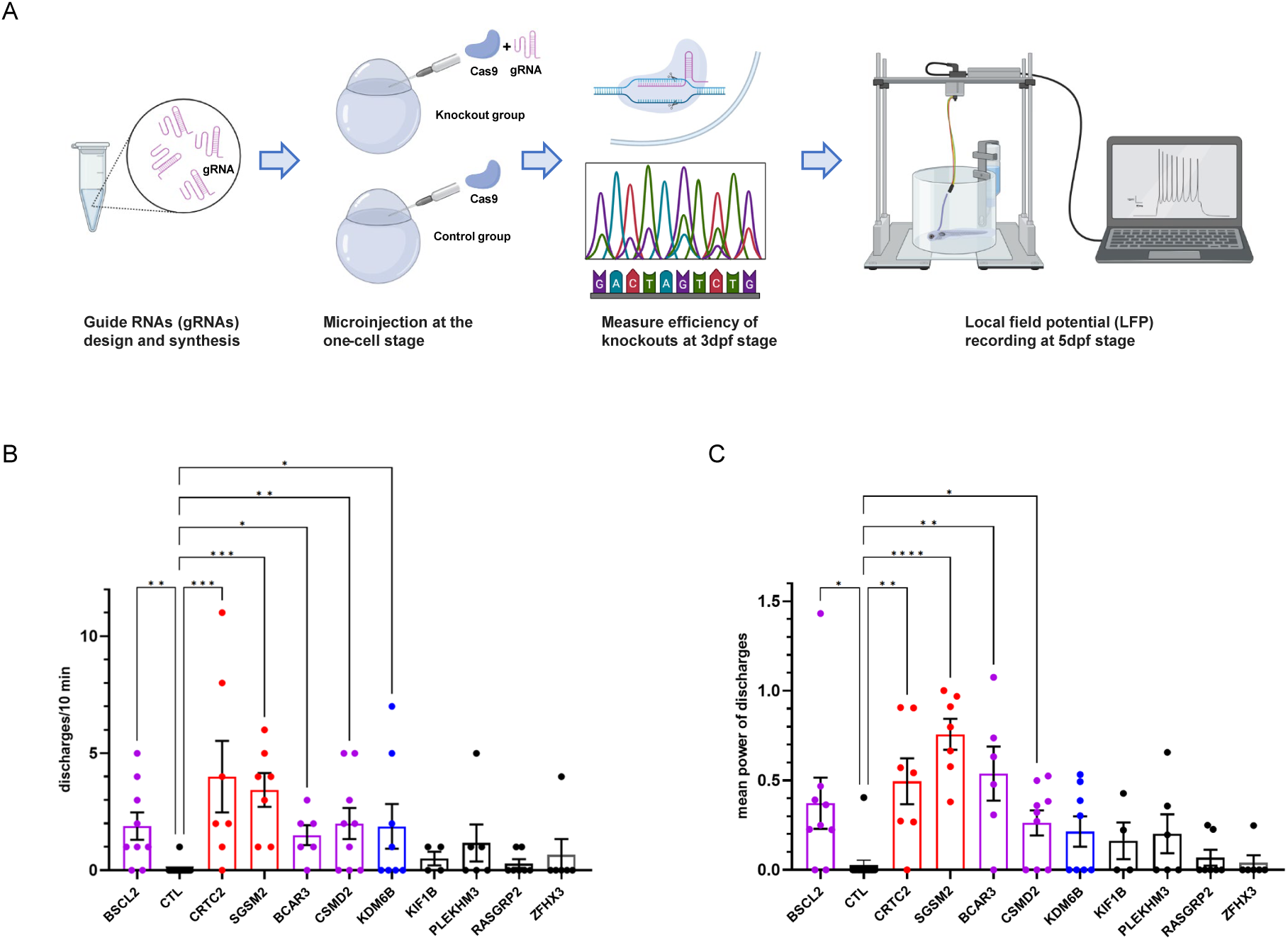
Rapid screening of novel epilepsy candidates based on Cas9-mediated gene knockout in F0 zebrafish. We observed that knocking out *CRTC2*, *SGSM2*, *BCAR3*, *CSMD2*, and *KDM6B* induced epileptic phenotypes in zebrafish based on local field potential recording. A: The scheme of candidate screening experiment using a zebrafish model. B: The X- and Y-axis indicate the name of candidates and numbers of discharges per 10 mins, respectively. C: The X- and Y-axis indicate the candidates’ names and the mean power of discharges, respectively. Statistical analyses were performed using the Kruskal–Wallis test. *: *P*-value < 0.01, **: *P*-value < 0.001, ***: *P*-value < 0.0001, ****: *P*-value < 0.00001.

We were able to conduct local field recordings in F0 knockouts of nine genes. The remaining six genes were excluded due to failure to generate specific gRNA (*CSMD1*, *PPP4R1*, and *SYNE1*, see **Supplementary Table 5** for details), gene disruptions that resulted in early lethality and thus made LPF recording impossible (*ADCY2* and *CDC45*), or a lack of expression in brain tissues (*CHRNG*)^47^. Compared with the controls, we observed significant increases (FDR-adjusted *P*-value < 0.05, Kruskal-Wallis Test) in synchronized epileptiform discharge (**Fig. 4B**) and duration when knocking out five novel candidates (*CRTC2*, *SGSM2*, *BCAR3*, *CSMD2*, and *KDM6B*) (**Fig. 4C**).

### Ca2+ imaging in HEK293T cells as cellular activity readout strategy

We further investigated whether overexpression of the constructs containing the variants in the novel candidates leads to a significant change in spontaneous calcium influx, which controls neuronal excitability and regulates calcium-sensitive intracellular signaling pathways^48–50^. Cellular activity based on calcium imaging in HEK293T cells has been widely utilized to investigate the function of genes or variants associated with neurological diseases^51–54^. To test the system, we first overexpressed the construct containing a pathogenic variant in a known epilepsy gene *BSCL2*^45,46^ in the HEK293T cells. We observed a more than two-fold increase in calcium fluctuation frequencies upon comparison of the vector containing the mutant allele versus the wild-type vector (**Figs. 5A-B**).

**Fig. 5:**
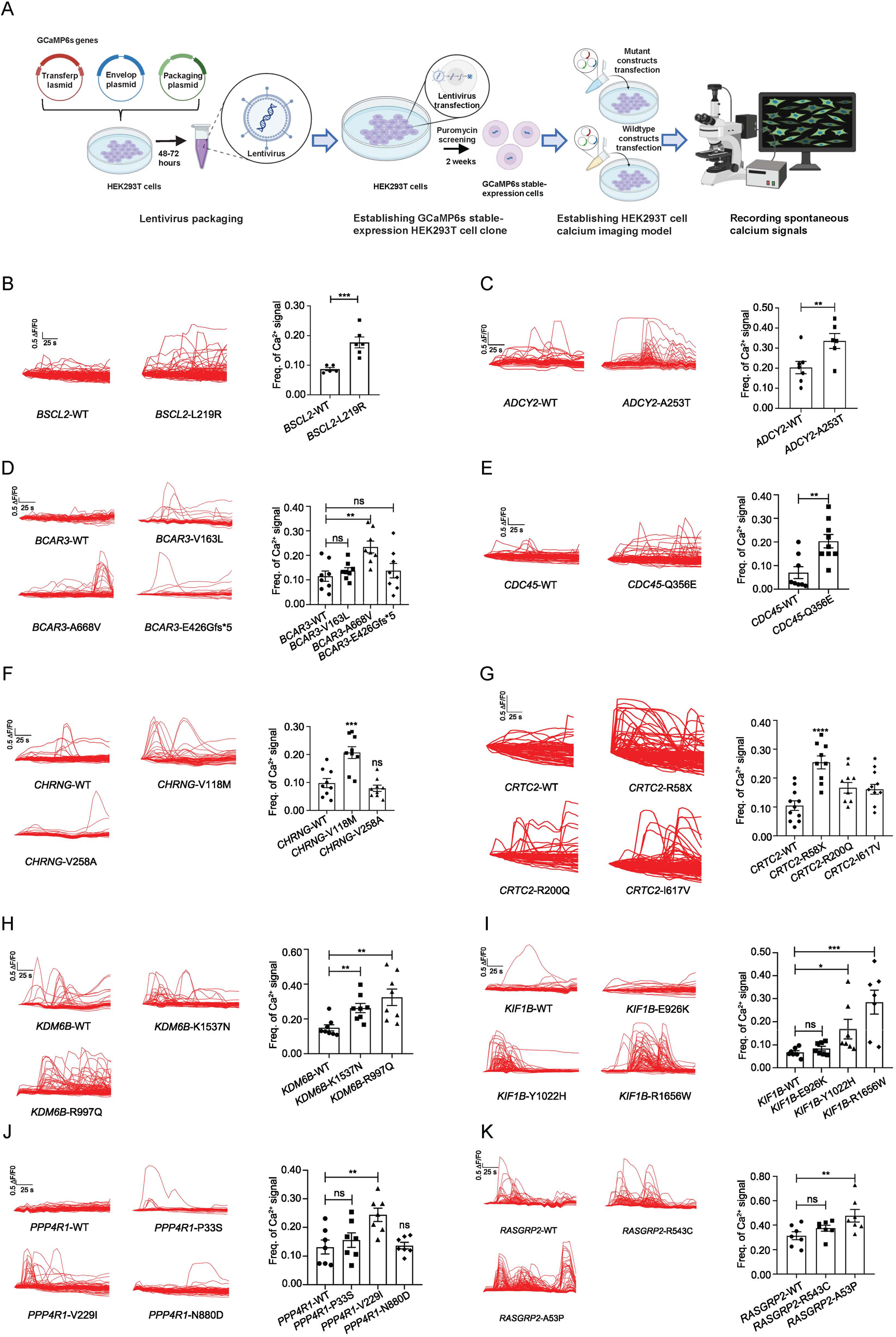
Rapid screening of variants in the 15 novel candidate genes for epilepsy was performed by assessing spontaneous calcium signals induced by overexpression experiment in human HEK293T cells. A: The scheme of rapid screening based on overexpression of the allele carrying the pathogenic variants identified in the novel candidates. B-K: Overexpression of the alleles harboring pathogenic variants in *BSCL2*, *ADCY2*, *BCAR3*, *CDC45*, *CHRNG*, *CRTC2*, *KDM6B*, *KIF1B*, *PPP4R1*, and *RASGRP2* resulted in a significant increase in spontaneous calcium signals. For each gene, the left panels depict representative calcium imaging traces and quantification of calcium influx frequency for the wild-type and variant alleles, while the right panels show the corresponding statistical analyses. The X-axis shows the variant name and the Y-axis is the frequency of the calcium signal. Statistical analyses were performed using the one-tailed unpaired t-test. *: *P*-value < 0.05, **: *P*-value < 0.01, ***: *P*-value < 0.001, ****: *P*-value < 0.0001. ns, not significant.

We excluded *CSMD1, CSMD2, SYNE1,* and *ZFHX3*, as their cDNAs are > 10,000 bps, which are too long to be cloned into the vector. A total of 33 vectors were constructed (including 11 vectors containing wild types and 22 vectors containing the mutants (**Supplementary Table 6**). We observed that the overexpression of 13 mutant constructs of nine genes (*ADCY2*, *BCAR3*, *CDC45, CHRNG*, *CRTC2, KDM6B*, *KIF1B, PPP4R1*, and *RASGRP2*) induced significant increases (*P*-value < 0.05, One-tailed unpaired *t-test*) in Ca2+ fluctuation frequencies compared with the wild-type constructs (**Figs. 5C-K**), suggesting elevated cellular activities. In particular, the findings from our overexpression experiments strongly suggest that the four candidates, such as *ADCY2, CDC45*, *CHRNG,* and *PPP4R1*, that can not be validated using zebrafish models are likely to have a pathological role in the context of human epilepsy.

### CRTC2

Out of the 15 candidates examined, *CTRC2* (also known as TORC2) stands out as one of the candidates demonstrating the greatest synchronized epileptiform discharge and prolonged duration in the zebrafish F0 knockout model (**Fig. 4**), along with the strongest elevated frequencies of Ca2+ fluctuations in our overexpression experiments **(Fig. 5)**. Previous studies have suggested that CRTC2 acts as a mediator of the mammalian target of rapamycin (mTOR) signaling pathway ^55^, which has been implicated in the pathogenesis of epilepsy ^56–60^. However, searches of the Online Mendelian Inheritance in Man (OMIM) database reveal no evidence for disease-causing genetic variants in *CRTC2* associated with epilepsy phenotypes.

In this study, we have discovered one *de novo* variant and two inherited ‘pathogenic’ missense variants in *CRTC2* across three unrelated probands. The *de novo* variant, c.172C > T, p.(R58X), results in the conversion of arginine to a premature stop codon in the TORC_N domain. This domain is thought to interact with the bZIP domain of CREB and the tetramer formation of TORCs^33^. Two inherited variants, c.599G > A; p.(R200Q) and c.1849A > G; p.(I617V), were found in the TORC_M and TORC_C domains, respectively. The function of TORC_M domain, which is situated between the N and C terminus of TORC proteins, is still unknown. TORC_C is negatively charged and bears a resemblance to transcription activation domains and thus it may play a role in the activation of transcription^34^. These variants were either absent (c.172C > T and c.1849A > G) or rare (with allele frequency < 0.0001, c.599G > A) in the samples of gnomAD and 1000 Genomes Project. Additionally, all six *in silico* programs in the present study predicted these three variants to have ‘deleterious’ effects.

Aligned with their functional importance on the molecular level, the most severe neurodevelopmental disorders phenotype was observed in the carrier (D285) of stop codon gain (R58X), exhibiting epilepsy, autism, and developmental delay, along with dysmorphic facial features such as a flat nose bridge and wide eye distance. In addition, D285 demonstrated evident symptoms of autism at early childhood and seizure onset at adolescent. Further, D285 developed a late-onset generalized tonic-clonic seizure without treatment with ASMs (**Supplementary Table 7**). In contrast, two missense variants carriers (D308 and D133) showed mild clinical phenotypes. Both had familial benign epilepsy with seizure onset at early childhood old and developed normally, with a family history of epilepsy or febrile seizures. D308 was diagnosed with GEFS+ with SE, and D133 was diagnosed with SeLECTS. Their seizures were controlled using clonazepam (D308) or oxcarbazepine (D133). Furthermore, cranial MRI revealed abnormal manifestations of arachnoid cysts in D285 and D133. The clinical characteristics of D133, D285, and D308 are summarized in **Supplementary Table 7**.

To further investigate functional connectivity across a range of temporal scales, we utilized a Maestro Pro multi-well microelectrode array (MEA) system (Axion BioSystems). This platform enables label-free assessment of neural activity and connectivity over time by recording the simultaneous spiking activity of hundreds of individual neurons (**Fig. 6A)**. This allows for functional electrophysiological characterization of both disease models and isogenic controls. Although the changes in the burst frequency (**Fig. 6F**, *P-*value = 0.056, One-way Anova test) and the number of burst (**Fig. 6G**, *P*-value = 0.072, One-way Anova test) did not reach statistical significance, we observed that overexpression of the *CRTC2* mutant in primary neurons using adeno-associated virus (AAV) led to significant increases in spike number (**Fig. 6B**, *P*-value = 0.045, One-way Anova test), mean firing rate (**Fig. 6C**, *P*-value = 0.0062, One-way Anova test), burst duration (**Fig. 6D**, *P*-value = 0.007, One-way Anova test), and the number of spikes per burst (**Fig. 6E**, *P*-value = 0.00032, One-way Anova test) compared to that of wildtype, indicating a state of hyperactivity in the mutant *CRTC2* overexpressing neurons, which aligns with our calcium imaging data in HEK293T cells (**Fig. 5**). We also performed electrophysiological recording of miniature excitatory postsynaptic currents (mEPSCs) of hippocampal neurons that overexpressing wild or mutant *CRTC2*. We observed that the overexpression of mutant *CRTC2* reduced the frequency of mEPSCs, but significantly increased the frequency (*P*-value = 0.039, One-way Anova test) and amplitude of mEPSCs (*P*-value = 0.00022, One-way Anova test), indicating a disruption of synaptic transmission in these neurons (**Figs. 6H-J**).

**Fig. 6:**
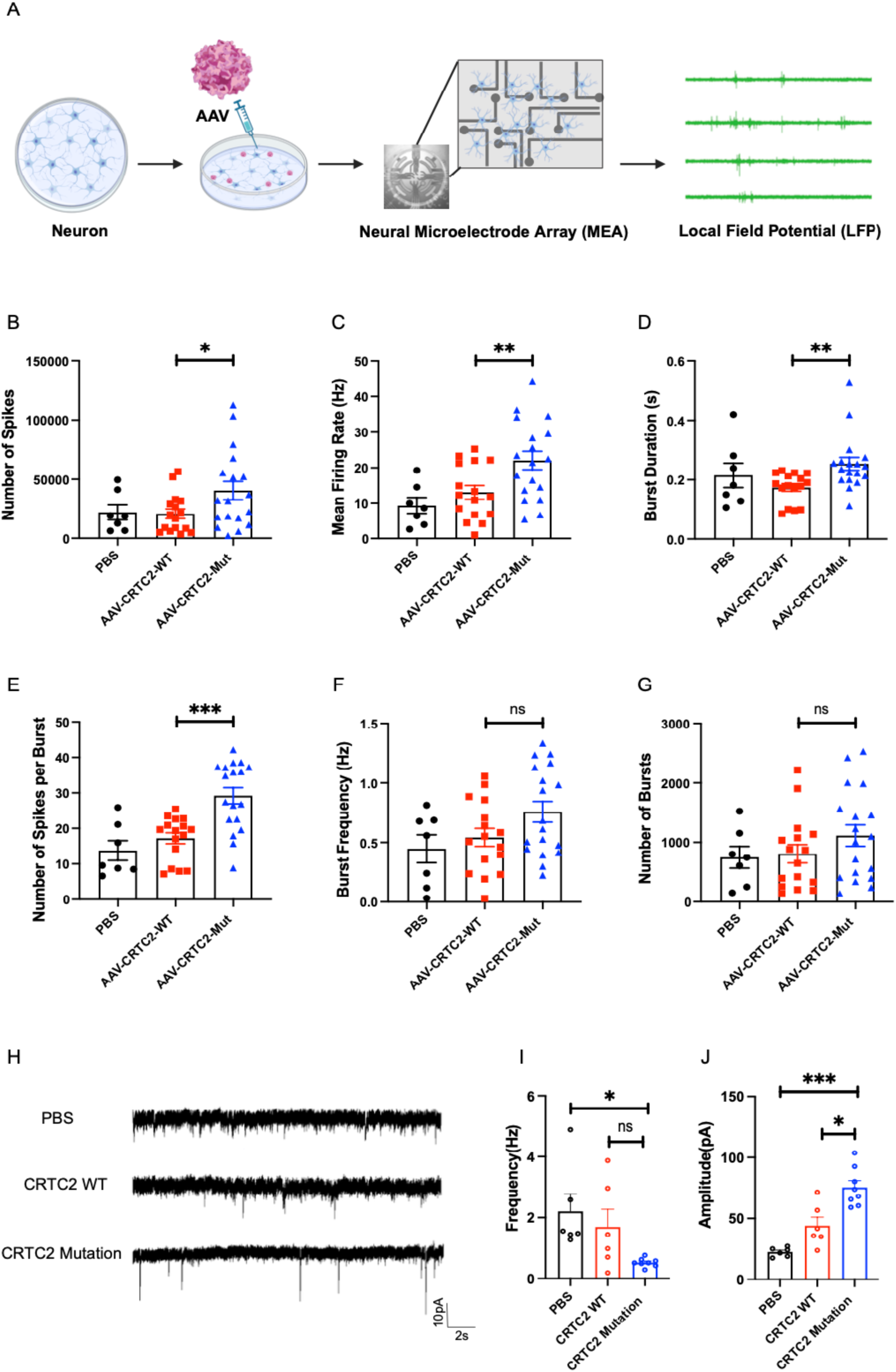
Overexpression of the mutant allele of *CRTC2* leads to cellular hyperactivity based on neural microelectrode array and patch clamp recording. A: A scheme of Neural microelectrode array experiment. Neuronal cells are cultured and then infected with AAV virus. Neural microelectrode array (MEA) devices were used to detect infected neurons, and the potential measured with MEA is called the local field potential (LFP). B-F: Overexpression of the mutant allele of CRTC2 leads to significant increase of number of spikes (B), mean firing rate (C), burst during per second (s) (D), number of spikes per burst (E) compared to wildtype. However, no significant differences were observed between burst frequency (F) and number of burst (G). H-J: Typical mEPSC recordings of hippocampal neurons treated with PBS, CRTC2 WT, and CRTC2 mutation. In (H), sample traces of mEPSC recordings are illustrated. Quantitative analyses of mEPSC frequency and amplitude are presented in (I) and (J), respectively. Scale bar: 10pA, 2s. mEPSC: miniature excitatory postsynaptic current. *: *P*-value < 0.05, **: *P*-value < 0.01, ***: *P*-value < 0.001. ns, not significant.

## Discussion

In this study, we analyzed the ES data of 963 children with epilepsy as well as their clinical phenotypes, treatments, and prognoses. We described the incidence and phenotypic variability of monogenic epilepsy, identified novel candidate genes, and reported prognoses of the patients based on their genetic etiologies.

Consistent with previous studies^10,29,61–63^, the patients in our cohort exhibited considerable heterogeneity in epilepsy types, age of seizure onset, seizure types, and genetic diagnoses (**Fig. 1A**). For instance, 67.9%, 30.2%, and 1.9% of patients had focal, generalized, and unknown seizure onset, respectively. Additionally, around 18% of patients displayed more than two seizure types (**Fig. 1B**). We also observed high incidences of neuropsychiatric comorbidities (64.0%), epileptogenic abnormalities (54.5%), and intractable epilepsy (37.7%). Furthermore, 43.8%, 13.0%, and 4.6% of patients had intellectual disability, ADHD, and ASD, respectively.

The genetic diagnosis rate in the present study is 39.6%, aligning with the rate reported in a prior large-cohort epilepsy based on ES^25,64–67^ and WGS data^29^. We observed a high burden of pathogenic variants in *SCN1A*, *KCNQ2*, and *DEPDC5* (**Fig. 1C**), which accounted for 21% of positive genetic diagnoses in our cohort. We also identified 26 CNVs in 40 patients and the duplications and deletions at 16p11.2 and 15q11.2 are the most frequent ones (**Fig. 1D**). However, prior studies showed that these CNVs are also common among both affected and unaffected populations and with low penetrance rates^68,69^. For example, the penetrance rate of 15q11.2 deletion is only 2.18% higher than the background risk^68^. Therefore, the pathogenicity of these CNVs still need to be evaluated in the future.

We observed a strong correlation between earlier seizure onset age and higher rates of positive genetic diagnosis. Specifically, the earliest seizure onset ages demonstrated the highest diagnosis rates. Additionally, higher diagnosis rates occurred in children with infantile-onset epilepsy and intellectual disability. This trend aligns with previous observations showing that early seizure onset in infancy correlates with a greater risk for developmental delays and poorer long-term outcomes^70–72^.

The comparison of genetic data across different cohorts, particularly between those of Chinese and European ancestries, provides novel insights into the distribution and impact of pathogenic or likely pathogenic variants within specific genes. In this analysis, several genes consistently stand out across various studies, such as *SCN1A* and *STXBP1*, which consistently rank among the top 15 genes harboring the highest burden of such variants. This suggests the important and widespread role of these loci in epilepsy pathogenesis across patients of diverse ancestries. However, the presence of discrepancies between cohorts of different ancestries highlights the complex interplay of genetic factors and population-specific genetic backgrounds. For example, genes like *GABRB3* and *DNM1* exhibit differences in their rankings between European and Chinese cohorts, where they are among the top 15 genes in European cohorts but not in Chinese cohorts. This observation underscores the importance of considering population-specific genetic variations and susceptibilities when interpreting genetic data and underscores the need for diverse representation in genetic studies to fully capture the spectrum of genetic variability.

Our analyses of ES data of the 582 patients who received negative clinical genetic diagnoses provide novel insights into the genetic etiology of pediatric epilepsy. In total, we identified 15 monogenic epilepsy candidates in > 2 unrelated patients. This provided genetic etiology for 34 (5.8%) probands who had negative clinical genetic tests.

We performed large-scale validation of the novel candidates using two complementary methods: CRISPR-Cas9 knockout in zebrafish and overexpression experiments in vitro. After excluding the genes unsuitable for these analyses, these two complementary methods suggest 11 genes can induce epileptic phenotypes in zebrafish or cell models (**Fig. 4 and 5**). For example, while knockout of *ADCY2* or *CDC45* resulted in early lethality in zebrafish, overexpressing vectors containing the variants identified in these two genes led to markedly increased calcium signaling frequencies (**Fig. 5**), indicating heightened cellular activity.

Although we were not able to validate *CSMD1, PLEKHM3, SYNE1, and ZFHX3* using cellular and zebrafish models, they have been implicated to involve in multiple neurological diseases. For example, variants in *CSMD1* and *CSMD2* have been implicated in autism spectrum disorder, and schizophrenia^73–77^*. PLEKHM3* is highly expressed in brain-related tissues (https://gtexportal.org/home/gene/PLEKHM3) and was suggested to play a role in adult brain cancer^78^. Mutations in *SYNE1* were associated with autosomal recessive arthrogryposis multiplex congenita 3 (myogenic type)^79^, autosomal dominant Emery-Dreifuss muscular dystrophy 4^80^, and autosomal recessive spinocerebellar ataxia 8^81^. *ZFHX3* was reported to be associated with autosomal dominant spinocerebellar ataxia 4^82^.

We reviewed the clinical phenotypes associated with the candidate genes in the present study in the OMIM database and previous publications. Including the aforementioned *CSMD1, CSMD2, PLEKHM3, SYNE1, and ZFHX3,* we observed some of the novel candidate genes identified in this study were associated with neurological and muscular diseases. For example, *KIF1B* was associated with autosomal dominant Charcot-Marie-Tooth disease (type 2A1)^83,84^ and with autosomal dominant neuroblastoma (type 1)^85^. Mutations in *KDM6B* were mainly associated with the autosomal dominant neurological phenotype of neurodevelopmental disorder with coarse facies and mild distal skeletal abnormalities^86^. Therefore, our analysis not only reveals the intricate interplay of genetic factors in the manifestation of various disorders, but also emphasizes the potential for shared pathways in the development of epilepsy and related neurological and muscular conditions.

We also observed that some of the validated candidates have not been reported to be associated with neurological diseases in the OMIM. For example, *RASGRP2*, was associated with platelet-type bleeding disorder-18^87^. *CHRNG* was associated with autosomal recessive Escobar syndrome^88^ and autosomal recessive multiple pterygium syndrome (lethal type)^89^. *CDC45* was implicated as pathogenic in the autosomal recessive disorders Meier-Gorlin syndrome 7^90^. The emergence of these candidates supports that our study has likely identified genes involving novel pathways not previously associated with epilepsy.

In summary, we performed extensive analyses of clinical and exome sequencing data from 963 patients. Our cohort exhibited substantial phenotypic heterogeneity in epilepsy types, seizure characteristics, and comorbidities. The overall diagnostic rate from exome sequencing was 39.6%, concurring with previous studies. We identified a significant burden of known pathogenic and newly characterized variants in established and novel candidate genes. Our findings expand the genotype-phenotype spectrum of pediatric epilepsy. This study also demonstrates the value of genetics-guided precision medicine, as 63.1% of the probands achieved seizure control using anti-seizure medications targeted to the molecular pathways implicated by their variants. Overall, our multi-level genomic analyses conducted in this study provide a comprehensive understanding of the genetic architecture underlying pediatric epilepsy, thereby offering valuable insights for enhancing diagnosis, prognosis, and personalized treatment strategies.

### The limitations of the present study

Our results greatly expand the genetic underpinnings of diverse epilepsy types, including DEE, GEE, and NAFE (**Fig. 3C**). While this study focused solely on pathogenic variants in more than two unrelated individuals with Mendelian inheritance patterns, further unraveling the genetic underpinnings of epilepsy necessitates accumulating patient genetic and clinical data, integrating more intricate inheritance patterns (e.g., oligogenic and polygenic), analyzing non-coding variants and repeat elements^91,92^, and leveraging large-scale association studies to elucidate the risk associated with common variants^17–21^.

## Material and Methods

### Participant recruitment and eligibility criteria

From January 1, 2016 to July 31, 2022, 963 pediatric patients with epilepsy was recruited at the Department of Neurology of the Children’s Hospital of Fudan University. All patients were diagnosed by neurologists based on multidimensional clinical information, including seizure type, electroencephalographic, and brain magnetic resonance imaging (MRI) results. To be eligible for inclusion, patients’ age of onset had to be under 18 years old, have a diagnosis of epilepsy based on the International League Against Epilepsy Criteria^93^, and have a suspicion of genetic cause. Patients with seizures that were likely caused by brain injuries, such as brain tumors, head trauma, hypoxic-ischemic injury, or infection of the central nervous system, were excluded based on their health records or questionnaires completed by doctors. Additionally, patients had to voluntarily accept genetic tests, and the IDs used in this study were not known to anyone outside the research group.

According to the International League Against Epilepsy (ILAE) Commission on Classification and Terminology guidelines formulated in 2017, seizure types are classified as focal onset, generalized onset, or unknown onset^4,94^. The ILAE Task Force on Nosology and Definitions 2022 methodology for epilepsy syndrome classification and definition categorizes syndromes based on typical phenotypes including age of onset, characteristic seizure types and electroencephalography (EEG) patterns, abnormal neuroimaging findings, and significant comorbidities^95^. For genetic epilepsies, we focused on the developmental and epileptic encephalopathy^6^ (DEE; MIM 308350), genetic generalized epilepsy (GGE; MIM 600669), and nocturnal frontal lobe epilepsy (NAFE; MIM 604364, 245570) phenotypic groups.

The study was approved by the ethics committee of the Children’s Hospital of Fudan University, and written informed consent was obtained from all parents or legal guardians according to the Declaration of Helsinki principles.

### Exome sequencing and analysis

Genomic DNA was extracted from the peripheral blood of the patients and their parents using QIAamp 96 DNA QIAcube HT Kit. Exome libraries were prepared using the SureSelect Human All Exon v5 Kit (Agilent Technologies, Santa Clara, CA, USA) or xGenExome V2 Kit (Integrated DNA Technologies, Coralville, IA, USA) in combination with the TruSeq Rapid PE (Paired-End) Cluster Kit and SBS Kit (Illumina, San Diego, CA, USA). The enriched libraries were sequenced using Illumina HiSeq 2000/2500 platform or DNBSEQ-T7 platform to generate paired-end 150 bp reads.

We trimmed the sequencing adaptors using Trimadap (version r11) (https://github.com/lh3/trimadap) and aligned the trimmed reads against the human reference genome (hg19) using BWA aligner^96^ with mem mode (version 0.7.15).

The mapping results were converted to the BAM format and sorted using SAMtools version 1.9^97^. We conducted variant calling using GATK toolkit^98^ (version 4.0.10.1) following the germline variant calling best practices. We used the variants with “PASS” flag in the downstream analyses.

We used ANNOVAR^99^ (version 2019-10-24) to annotate the variants and to predict their functional impacts based on a set of *in silico* pathogenicity prediction programs, including SIFT^100^, PolyPhen-2^101^, MutationTaster^102^, MutationAssessor^103^, FATHMM^104^, FATHMM_MKL^105^, fitCons^106^, CADD^107^, and DANN tools^108^.

The kin relationship of each trio was confirmed using Plink v1.90b5.21^109^.

We defined a variant as “pathogenic” when it causes gene disruption (e.g., mutations at splicing sites, pre-stop codon gain and stop codon loss, start codon loss, open reading frame shift) or damaging missense effect (predicted to have deleterious effects by at least 3 in silico programs or with CADD score greater than or equal to 20 or DANN score greater than or equal to 0.93) and with frequency ≤ 0.001 in the EAS population of gnomAD, 1000 Genomes Project, and ExAc databases.

We then classified *de novo* or recessive variants based on co-segregation analyses. Additionally, we assessed whether these filtered variants were located in the known epilepsy genes in the Online Mendelian Inheritance in Man (OMIM, https://www.omim.org/), HGMD^36^, Clinvar^37^, and PubMed databases. The unreported mutation was subsequently categorized as pathogenic, likely pathogenic, uncertain significance, or negative, following the American College of Medical Genetics and Genomics (ACMG) Guidelines^110,111^.

Any genes that contained variants that survived our filtering and interpretation procedures were considered as novel candidate genes of epilepsy. We further prioritized candidate genes that were identified in at least two unrelated probands.

### A comparison with the Epi25 dataset

We obtained the burden test results between 20,979 patients with epilepsy and 33,444 controls by Epi25 Collaborative from https://storage.googleapis.com/exome-results-browsers-public/downloads/2022-12-01/Epi25/Epi25_gene_results.tsv.bgz. We converted the gene symbols of the 15 candidate genes to ENSEMBL ID using bioDBnet (https://biodbnet-abcc.ncifcrf.gov/db/db2db.php). We then extracted the gene-based burden test results of damaging missense ultra-rare variants for the candidate genes and used the exome-wide significance threshold to determine if the candidate genes are significant in this large-scale genetic study^15^ (**Supplementary Table 2)**.

### Plasmid constructs

To obtain the wild-type of candidate genes, we subcloned complementary DNA (cDNA from Hela cells) or plasmid templates (**Supplementary Table 6**). We amplified PCR reactions as per the KOD OneTM PCR Master Mix (Code No.TAK-101, TAKARA) guidelines, and the purified PCR products were then constructed into a BamHI-HF (Catalog#: R3136L, NEB) digested FUGW-2*His-P2A-mCherry vector, which was based on the FUGW vector and modified by our lab, using the ClonExpress® II One Step Cloning Kit (Catalog#: C112-02, Vazyme). The missense or stop-gain mutations corresponding to each candidate gene were generated through site-directed mutagenesis using the M5 HiPer Site-Directed Mutagenesis Kit (Catalog#: MF129-02, Mei5 Biotechnology, Co., Ltd). A list of the primers used can be found in **Supplementary Table 9**.

### Zebrafish strains

Our animal use protocol was approved by the Fudan University Shanghai Medical College Institution Animal Care and Use Committee (20210302-149), and all animals were handled in strict compliance with the Fudan University Regulations on Animal Experiments.

To maintain the zebrafish, we followed standard protocols^112^ and used the AB strain for this study. Zebrafish larvae were obtained through natural spawning and were raised under 14 h/10 h light/dark daily cycles at 28.5 °C.

### Generation of F0 knockouts

To generate F0 knockouts in zebrafish, we followed a published method^43^ (Kroll et al., 2021). We designed three guide RNAs (gRNAs) per candidate (**Supplementary Table 5**) and injected them with Cas9-gRNA RNP mixtures at the one-cell stage. For the control group, zebrafish were injected with Cas9 protein only. The efficiency of knockouts was measured using TIDE^113^ and DECODR^114^.

### Local field potential (LFP) recording

LFP recordings were conducted following the procedures from previously published studies with some modifications^44^. Specifically, each zebrafish larva at 5-6 days post-fertilization (dpf) was mounted in 1% low melt point agarose. A recording electrode was then inserted into the dorsal surface of the forebrain, while a reference electrode was placed near the surface of the hindbrain. Electrodes were insulated stainless steel, and the recording conditions were the same as those described in prior work.^115^ LFP signals were sampled at 5 kHz, denoised via wavelet decomposition, and any epileptiform discharges were identified and quantified using custom MATLAB scripts (https://github.com/gpeng1903/danioLFP).

### Screening base on spontaneous calcium signals in HEK293T cells

The plasmid pLVX encoding calcium indicator GCaMP6s gene was utilized to transfect HEK293T cells with the aid of Lipofectamine 2000 (Invitrogen) and Lentivirus packaging components. Virus particles were collected and purified from the culture medium after 48-72 hours of transfection and concentrated using a lentivirus purification kit (Yeasen, 41101ES50). For the generation of stable cell lines overexpressing GCaMP6s, 2 μL of purified lentivirus was added to HEK293T cells and puromycin was employed to select a single clone. The cells were grown in puromycin-containing DMEM for two weeks, with the medium changed every two days until a stable clone was established. To prepare for the experiment, GCaMP6s stable-expression HEK293T cells were seeded onto 35 mm Mattek dishes with coverslips. Wildtype or mutant constructs were then transfected into the cells using Lipofectamine 2000. After a further 48 hours of culture, spontaneous calcium signals were recorded every 5 seconds using an Axio Observer.Z1 microscope (Zeiss). Fluorescence intensity of GCaMP6s was quantified using ImageJ (NIH). The baseline of 5 seconds before the peak signal was used as F0, and only △F/F0 that was more than 2SD of F0 was recognized as a peak calcium influx.

### Multielectrode array (MEA) experiment

Primary hippocampal neurons were dissociated and cultured in 24-well MEA plates. On day in vitro (DIV) 3, we added adeno-associated virus (AAV) to overexpress either wildtype or mutant *CRTC2* alleles into the neurons. On DIV 7, we exchanged the cell culture medium. MEA recordings were then carried out between DIV 13-15.

### Electrophysiology and data analysis

Hippocampal pyramidal neurons were selected for whole-cell patch-clamp recordings based on their morphology on an inverted microscope (Leica, DM-IRB). Data were collected with an Axopatch-700B amplifier, a Digidata-1440A digitizer and pClamp 10.2 software (Molecular Devices), low-pass filtered at 2 kHz, and digitally sampled at 10 kHz. Patch pipettes (4.0 - 6.0 MΩ) were pulled from borosilicate glass (Sutter Instruments) and filled with one of two types of pipette solution. For miniature EPSCs (mEPSC) recordings, the pipette solution contained the following (in mM): 120.0 Cs-methanesulfonate, 0.6 EGTA, 2.8 NaCl, 5.0 MgCl2, 2.0 ATP, 0.3 GTP, 20.0 HEPES, and 5.0 QX-314, and adjusted to pH 7.2 with CsOH (305-310 mOsm). The standard extracellular solution contained the following (in mM): 119.0 NaCl, 2.5 KCl, 2.0 CaCl2, 2.0 MgCl2, 25.0 HEPES, and 30.0 D-glucose. The pH was adjusted to 7.4 with NaOH. mEPSCs was recorded in the presence of both 10 μM bicuculline and 1 μM tetrodotoxin (TTX; Abcam) in the extracellular solution. Recordings were made at a holding potential of −70 mV. QX-314 was added to the pipette solution to block the GABAB-mediated currents and to prevent the generation of Na-dependent action potentials. QX-314 and bicuculline were purchased from Tocris Bioscience. mEPSC was analyzed offline using Clampfit 10.7 (Molecular Devices) and Igor 5.3 (Wavemetrics). Recordings with series resistance 20 MΩ were excluded from analysis. Synaptic events 5 pA were detected after creating a unique template for each neuron. All recordings were performed at room temperature (25°C). All data are presented as mean±SEM. Investigators were blinded to the treatments of the cells during experiments.

### Statistical analysis

GraphPad Prism 9 and Matlab were used to analyze and plot LFP recording data. The Kruskal-Wallis test was used to examine whether the differences between the knockouts and control reached statistical significance or not. The multiple comparisons were corrected by controlling the false discovery rate using the BKY method.

## Data availability

Data supporting this study are available from at the National Genomics Data Center (https://ngdc.cncb.ac.cn) with accession number HRA006570. Access to the data is subject to approval and a data sharing agreement due to IRB restrictions.

## Table captions

**Supplementary Table 1**. The demographic and clinical characteristics of 963 patients in the present study.

**Supplementary Table 2**. Burden test results of 15 candidate genes identified in the present study in the Epi25K dataset^15^.

**Supplementary Table 3**. KEGG enrichment results using 15 candidate genes in the present study and 978 known epilepsy genes in the Genes4Epilepsy database.

**Supplementary Table 4**. GO and KEGG enrichment of the genes with the pathogenic *de novo* variants that were identified in individual samples.

**Supplementary Table 5**. Guide RNAs (gRNAs) that were used to generate F0 zebrafish knockouts.

**Supplementary Table 6**. Plasmid constructs that were used in the overexpression experiments in HEK293T cells. We constructed 33 plasmids including 11 for wide type (WT) and 22 for variants.

**Supplementary Table 7**. The clinical characteristics of three patients carrying pathogenic mutations in *CRTC2*.

**Supplementary Table 8**. The clinical characteristics of the patients carrying novel epilepsy candidate genes.

**Supplementary Table 9**. The primer sequences for constructing plasmids of novel epilepsy candidates. F:forward primer, R: reverse primer.

